# A central research portal for mining pancreatic clinical and molecular datasets and accessing biobanked samples

**DOI:** 10.1101/2024.07.25.24309825

**Authors:** Jorge Oscanoa, Helen Ross-Adams, Abu Z. M. Dayem Ullah, Trupti S. Kolvekar, Lavanya Sivapalan, Emanuela Gadaleta, Graeme J. Thorn, Maryam Abdollahyan, Ahmet Imrali, Amina Saad, Rhiannon Roberts, Christine Hughes, PCRFTB, Hemant M. Kocher, Claude Chelala

**Author notes:** Corresponding Author: Prof C Chelala. These authors contributed equally (Jorge Oscanoa, Helen Ross-Adams).

## Abstract

The Pancreas Genome Phenome Atlas (PGPA) is dedicated to the analysis of pancreatic datasets from four primary sources (Cancer Genome Atlas, International Cancer Genome Consortium, Cancer Cell Line Encyclopaedia, Genomics Evidence Neoplasia Information Exchange) that together form the foundation of -omics profiling of pancreatic malignancies and related lesions (n=7,760 specimens). Multiple user-friendly analytical tools to explore the associated molecular data from these primary specimens and cell lines are available. Crucially, PGPA is the access point for Pancreatic Cancer Research Fund Tissue Bank – the only national pancreatic cancer biobank in the UK, and will facilitate effective sharing of multi-modal molecular, histopathology and imaging data from biobank samples (>60,000 specimens from >3,400 cases and controls; 2,037 H&E images from 349 donors) and accelerate validation of *in silico* findings in patient-derived material. This places PGPA at the forefront of biomarker-based research, providing the user community with a distinct resource to facilitate hypothesis-testing on public data, validate novel research findings, and access curated, high-quality patient tissues for translational research. To demonstrate the practical utility of PGPA, we investigate somatic variants associated with established transcriptomic subtypes and disease prognosis: several patient-specific variants are clinically actionable and may be leveraged for precision medicine.

## Introduction

Pancreatic ductal adenocarcinoma (PDAC) is predicted to become the second leading cause of cancer-related mortality worldwide before 2040(1). It has dismal 5-year survival rates of 3-15%(1, 2), largely due to late disease detection and few effective treatment options. Alarmingly, the incidence of early-onset pancreatic cancer is also increasing, in contrast to most other solid tumours(3). Better tools for patient stratification and treatment response are thus essential to improve survival outcomes. However, the pancreatic cancer research community is relatively small – investigators tend to develop bespoke collections of samples that may be unusable beyond the breadth and scope of their ethical approval and storage conditions. Sample collection protocols also vary widely, further limiting the translation of derived results to clinical benefits.

Most existing biomarkers for monitoring treatment or assessing prognosis are not based on the molecular attributes of PDAC tumours and have shown limited sensitivity and/or specificity in prospective settings(4). Numerous studies into the genomic and transcriptomic determinants of tumour development and progression are available, but their findings are dispersed across multiple resources, and can be difficult to access and translate into meaningful survival or treatment benefits for patients by those without computational expertise. This highlights the pressing need for simplified, integrated data mining and analysis tools to improve accessibility to clinical and molecular information from disparate sources and enable laboratory and clinical researchers to easily and effectively cross-query large multi-omics datasets, to fuel new discoveries in pancreatic diseases.

Here, we present the Pancreas Genome Phenome Atlas ([PGPA](https://pgpa.bcc.qmul.ac.uk/home/)), an intuitive, online portal that links numerous multi-modal datasets to an active biobank where users can both validate their findings, and/or apply for samples to confirm *in silico* findings. This represents a significant expansion in functionality from our old 2018 Pancreas Expression Database^8^ (now retired): more analytic tools, expanded datasets to encompass the broad spectrum of pancreatic lesions enriched with multi-modal data (demographic, clinical, transcriptomic and genomic) and links to an active biobank. This website is free and open to all users and there is no login requirement.

To remain abreast of the evolving nature of integrative multi-omics workflows, we have included a broad range of available datasets and placed this essential resource at the centre of an established framework (Table 1).

**Table 1.** Summary of the new features in PGPA.

| Features | PED<br>(archived)<sup>8</sup> | PGPA |
| --- | --- | --- |
| **The Analytics Hub** |  |  |
| **Publicly available data sources (pancreas-specific)** |  |  |
| PubMed<sup>a</sup> | X |  |
| TCGA<sup>b</sup> | X | X |
| ICGC |  | X |
| GENIE<sup>c</sup> | X | X |
| CCLE<sup>d</sup> | X | X |
| **Analytical features** |  |  |
| Principal components analysis | X | X |
| Gene expression profiles | X | X |
| Correlation analyses | X | X |
| Gene networks | X | X |
| Survival analyses<sup>e</sup> | X | X |
| Variant identification | X | X |
| Somatic gene interactions | X | X |
| Reactome & oncogenic pathway analyses |  | X |
| Improved clinical annotations to visualize & query publicly available datasets |  | X |
| MAFtools genomic analyses and summary visualisations |  | X |
| Tumour mutational burden |  | X |
| Clinically actionable genes/proteins & associated drugs |  | X |
| Cohort comparison by clinical/molecular feature |  | X |
| Gene intersections between filtered datasets |  | X |
| **The PCRFTB Data Module** |  |  |
| Data return module to host both -omics and experimental datasets |  | X |
| Integrated primary/secondary care clinical data |  | X |
| Apply for samples |  | X |

<sup>a</sup> Less-used literature mining module
<sup>b</sup> Includes essential filters based on cancer subtype
<sup>c</sup> Includes full set of 6,633 pancreatic cancers of all types, with additional clinical and molecular information.
<sup>d</sup> Somatic variant dataset; includes the full set of 60 primary and metastatic tumour derived cell lines.
<sup>e</sup> Includes analyses based on mutational status and mRNA level

PGPA is also integrated as the major bioinformatics platform of the UK’s national Pancreatic Cancer Research Fund Tissue Bank (PCRFTB), to facilitate investigative biomarker-based research and data sharing between clinicians and scientists. This is powered by a customised version of [SNPNexus](https://www.snp-nexus.org/v4/), a versatile platform for the functional annotation of known and novel sequence variation(5), designed to reduce the analytical burden associated with large-scale genomic datasets and facilitate the straightforward identification of biologically and clinically relevant genetic variants in patients.

The PCRFTB is the world’s first national pancreas tissue bank and has been collecting blood, urine, saliva and solid tissue samples from patients recruited at nine participating centres across the UK NHS since 2015, making it a valuable resource for translational research. Tissues are available from patients with pancreatic and hepatobiliary diseases, including resectable and unresectable cancer. Blood, urine and saliva samples from patients, their first-degree relatives and other healthy volunteers are also available, as well as cancer organoids and cancer-associated fibroblasts. (Figure 1). These are accompanied by extensive, verified clinical, histological and imaging data that is continually updated – median 300 data points per visit, with some donors providing longitudinal samples at multiple visits throughout their treatment journey. Best practice and ongoing technical research ensures available biological materials are of high quality, to support reliable and reproducible results(6, 7) In addition to samples, digitised radiological images are available for 171 patients with malignant, pre-malignant and benign pancreatic diagnoses, and 2,037 H&E images from 349 donors are also currently available.

**Figure 1.**
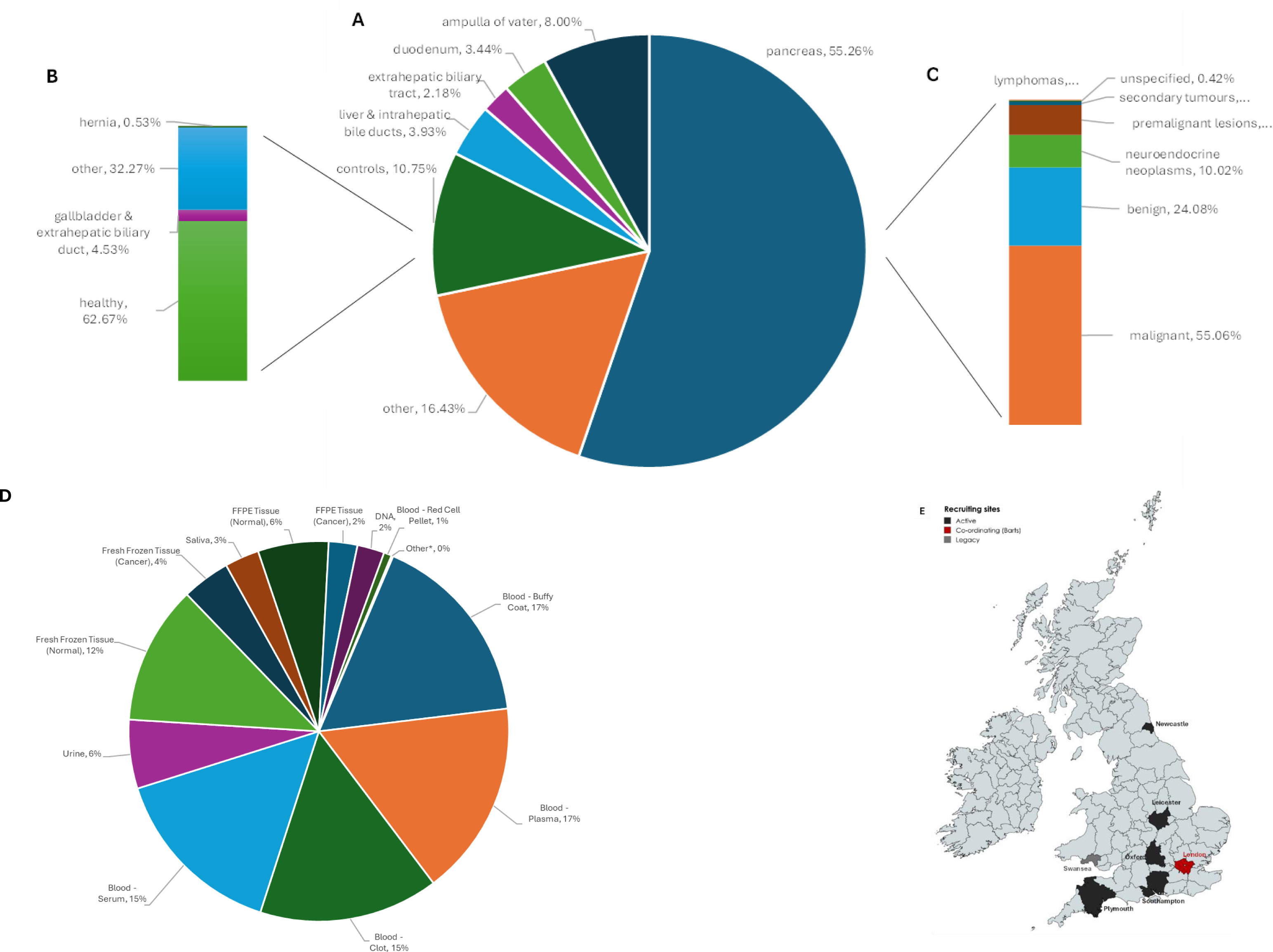
Summary of available tissue types. (**A**) The proportion of >3 400 unique organ site tissues available for study in the UK national Pancreatic Cancer Research Fund Tissue Bank, with the breakdown of controls (**B**) and pancreas (**C**) highlighted. (**D**) Distribution of >60 000 [PCRFTB](https://www.thepancreastissuebank.org/) specimens by type, across all patients. Details are updated weekly. Additionally, radiological imaging is available for 171 patients with malignant, pre-malignant and benign pancreatic diagnoses, and >2,000 H&E images from 349 donors. Samples can be applied for [here](https://pcrftbtissueaccess.bcc.qmul.ac.uk/). (**E**) Geographical locations of PCRFTB patient recruitment sites. Map created with mapchart.net. *=pancreatic juice, CTC, bile, organoids.

The PCRFTB is further supported by a Data Return policy to maximise the use of available samples by linking each patient/donor with an enriched ‘digital fingerprint’ encompassing molecular, transcriptomic, proteomic, imaging and longitudinal clinical data. All donors provide written, informed consent, and all samples are collected, processed and stored at each of the participating centres (Barts, Leicester, Swansea, Oxford, Royal Free (London), Southampton, Newcastle, Plymouth, The London Clinic) under one Research Ethics Committee reference (13/SC/0593, renewed 18/SC/0629, renewed 23/SC/0282) and using standardised protocols, quality assurance and quality control policies ensuring consistency across the collection.

Via PGPA, researchers can directly query and apply to PCRFTB for samples, specimens and/or imaging data that match user-determined criteria. A link to the PCRFTB Tissue Request System allows users to submit Expressions of Interest directly to the Tissue Bank using an online application form.

By providing a dynamic hub for the analysis of publicly available pancreatic datasets and ongoing research data generated from biobanked samples, PGPA allows researchers to access a broad range of pancreas-specific molecular information freely and quickly. The flexibility of this hub allows molecular alterations with biological and clinical relevance to be identified and prioritised for downstream validation.

## The Analytics Hub

The web-based Analytics Hub includes a broad range of pancreas-related, publicly available -omics datasets, together with extensive analytical features and visualisation options (Table 1).

### Publicly available data sources

PGPA analytics hub hosts publicly available clinical and molecular datasets from four core sources: The Cancer Genome Atlas (TCGA)(8) whole exome sequencing (WES) data, with filters for all available pancreatic cancer types; the Cancer Cell Line Encyclopaedia (CCLE)(9), including somatic mutation and mRNA expression data for the complete set of 60 pancreas cell lines from primary and metastatic tumours; the Genomics Evidence Neoplasia Information Exchange (GENIE)(10) v13.0, including simple somatic mutations and clinical data from the *complete* set of 6,633 patients with pancreatic cancer of any type; and the now archived complete International Cancer Genome Consortium (ICGC)(11, 12) dataset, including whole genome and RNA sequencing data from both adenocarcinoma (PACA-AU; PACA-CA) and neuro/endocrine tissues (PAEN-IT; PAEN-AU). These sources host data generated by both national and international consortia efforts to sequence and analyse cancer genomes and biology, including pancreatic malignancies. Analysed and quality-controlled data files were downloaded from the respective sources and used without further processing. PGPA 2024 uses the most recent data releases, including linked clinical data when available.

### Advanced filtering of public datasets

Public datasets may be queried according to the clinical characteristics of each study cohort (Figure 2A). Filtering options have been selected based on relevance to disease development and pathogenesis, and the depth of annotation provided in available clinical data for each cohort. Implemented filters are accompanied by dedicated visualizations of clinical summaries for each respective study cohort (Figure 2B). These include filters based on patient-related factors (cancer type, sex, age, diabetes, family history, ethnicity, survival; Figure 2C), and tumour characteristics (stage, grade, *KRAS* somatic mutation status) (Figure 2D), which allow for trends in data to be clearly observed: e.g. survival beyond 3 years is very low for PDAC compared to neuroendocrine tumours (Figure 2B). Crucially, it is possible to filter each dataset by diagnosis, allowing researchers to focus on different pancreatic lesions individually (e.g. IPMN, ductal adenocarcinoma, neuroendocrine, adenosquamous, mucinous) that have different molecular alterations and clinical prospects, since using unstratified sample sets has been shown to yield unreliable results.(13, 14)

**Figure 2.**
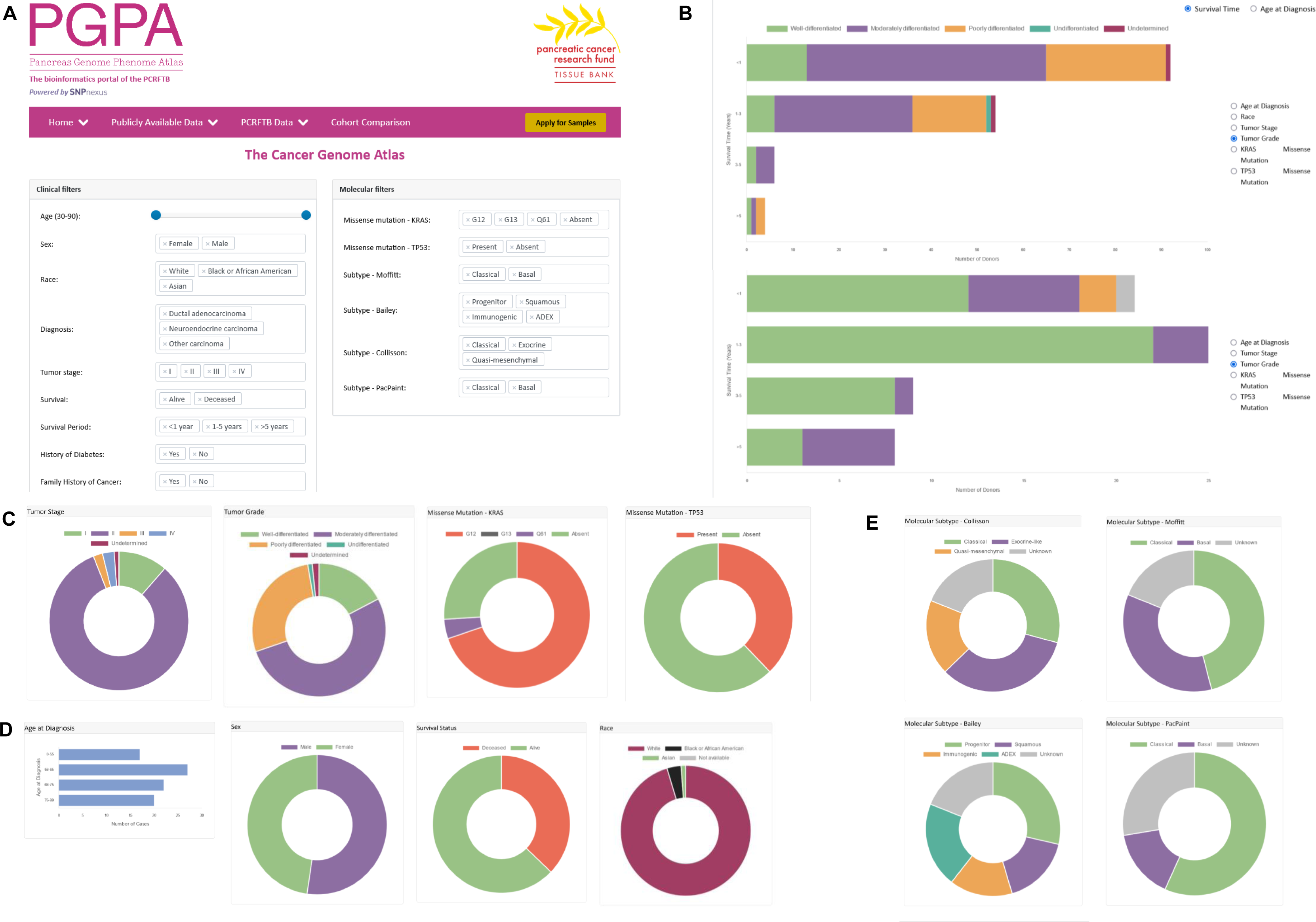
Advanced filtering options and clinical summaries for publicly available PDAC datasets. **(A)** Available data can be filtered according to various patient-related factors and tumour characteristics, including the stratification and analysis of cohorts according to KRAS and TP53 mutational status and established transcriptomic (TCGA, ICGC), genomic (ICGC) or histologically-derived AI (PacPaint) subtypes. **(B)** Dynamic bar charts allow multiple covariates to be viewed in relation to each other: e.g. survival trends in PDAC (TCGA) and neuroendocrine (ICGC PAEN-AU). **(C, D, E)** Each filtered attribute can be visualized as clinical summaries for each study cohort. Alternatively, data can be downloaded as .csv or .xls files, for offline analysis.

### Characterising the genomic characteristics of established PDAC molecular subtypes

PGPA facilitates the stratification and analysis of TCGA and ICGC cohorts according to their molecular subtype classifications, as determined by hallmark transcriptomic(15–17) and genomic (ICGC only)(18) studies, and recent histopathology-based artificial intelligence (AI) predictions in matched TCGA samples(19)(Figure 2E). We demonstrate the implementation of this clinically useful feature below.

Collisson et al. (2011) originally identified 3 subclasses of PDAC tumours with different clinical outcomes and treatment responses, termed *quasi-mesenchymal* (QM) (worst prognosis), *classical* (best prognosis) and *exocrine-like*, using hybridisation array-based mRNA expression data from primary untreated resected PDAC(15). Next, Moffitt et al (2015) analysed bulk tumour tissues from treatment-naïve primary resected PDAC tumours using virtual microdissection to exclude transcripts native to the normal pancreas and the tumour microenvironment, and reported 2 distinct tumour subtypes (*basal* and *classical*) as well as 2 classifications based on peritumoural stromal tissues (*activated* and *normal*)(16). *Basal* subtype tumours were associated with a poorer overall patient survival compared to *classical* tumours, which overlap significantly with the Collisson *classical* subtype. Subsequently, Bailey et al. performed RNA-sequencing of bulk primary untreated resected tumour tissues from 328 PDAC tumours and resolved four stable tumour classes (*squamous*, *pancreatic progenitor*, *immunogenic* and *aberrantly differentiated endocrine exocrine* (ADEX)), each governed through the differential expression of transcription factors and their targets involved in lineage specification during pancreatic development(17). *Squamous* subtype tumours overlapped with previous *basal* (Moffitt) and *QM* (Collisson) classifications and were associated with the poorest overall prognosis in patients. Recent investigations of these proposed classifications have corroborated the presence of 2 overarching transcriptomic subtypes of PDAC tumours comprising *basal-like/squamous* and *classical/progenitor* that have shown relevance for defining survival outcomes in patients, with remaining subtypes (*exocrine-like, ADEX*) shown to have confounding associations with poor tumour cellularity(20, 21). Most recently, Saillard et al (2023) used an artificial intelligence model trained and validated on 5 independent surgical and biopsy cohorts with RNAseq and histology data (n=598), including n=126 TCGA samples to further refine these tumour subtypes(19). This approach recapitulated the known *basal/classical*^17^ tumour subtypes at the whole H&E slide level but detected variable proportions of basal cells in samples previously categorised as classical subtype when slides were analysed at 112 <u>µ</u>m tile size level. This changed survival outcomes in 39% of cases classed as *classical* subtype by bulk RNAseq analysis, with the impact apparently proportional to the percentage basal cell content.(19)

At the DNA level, whole genome sequencing (WGS) and copy number variant (CNV) analysis performed on 100 treatment-naïve, macro-dissected PDAC tissues (ICGC Australia) identified four disease subtypes with distinct patterns of structural variation (*scattered, locally rearranged, stable, unstable)* and clinical utility, with *unstable* subtype characterised by a very high degree of genomic instability throughout the genome and encompassing defects in DNA damage repair (DDR) pathways that confer susceptibility to PARP inhibition (PARPi) and/or platinum chemotherapies(18).

### Application of subtype-specific filtering criteria in study cohorts

Subtype-specific characteristics can be explored using TCGA (n=185) and ICGC Australia (PACA-AU, n=461) and Canada (PACA-CA, n=317) pancreatic cancer cohorts. Molecular subtype classifications according to Collisson(15), Moffitt(16), Bailey(17) and/or Saillard(19) are available for n=134 of the 156 confirmed PDAC patients included in the TCGA cohort(20). Alternatively, ICGC-AU cohorts can be analysed according to the subtype classifications proposed by Bailey et al (2016)(17) (n=95 patients total; 81 PDAC) and/or Waddell et al (2015)(n=86 patients total; 85 PDAC).(18) Given the prognostic relevance of these transcriptomic subtypes, here we explore the associated genomic features of TCGA PDAC tumours classed unanimously as either classical/progenitor (n=27) or QM/basal-like/squamous (n=16) by all three subtyping systems (Collisson/Moffit/Bailey), as an example of PGPA Analytics Hub.

### Subtype-specific somatic variations

Comparisons between the genomic characteristics of each TCGA subtype showed different gene sets mutated in *classical* and *basal* subtypes (Figure 3A, B). This was also true for PACA-AU prognostic subtypes (progenitor vs squamous) (Supplementary Figure 1A, B), but with little consensus between the two cohorts (Supplementary Figure 1C). To more reliably identify subtype-specific genetic variations robust to inevitable inter-study variability (e.g. tissue heterogeneity; WES vs WGS(22)), we considered the *union* of the top 25 most frequently mutated genes between similar prognostic groups for the two largest datasets (TCGA+ICGC). This revealed a handful of common genes detected at >10% prevalence (*KRAS, TP53, CDKN2A, MUC16, LRP1B, AFF2, FAT4*), but with most genes/variants being subtype-specific (Figure 3C), consistent with (1) the early acquisition of these common alterations during PDAC tumour development and (2) the molecular heterogeneity of PDAC tumours(20).

**Figure 3.**
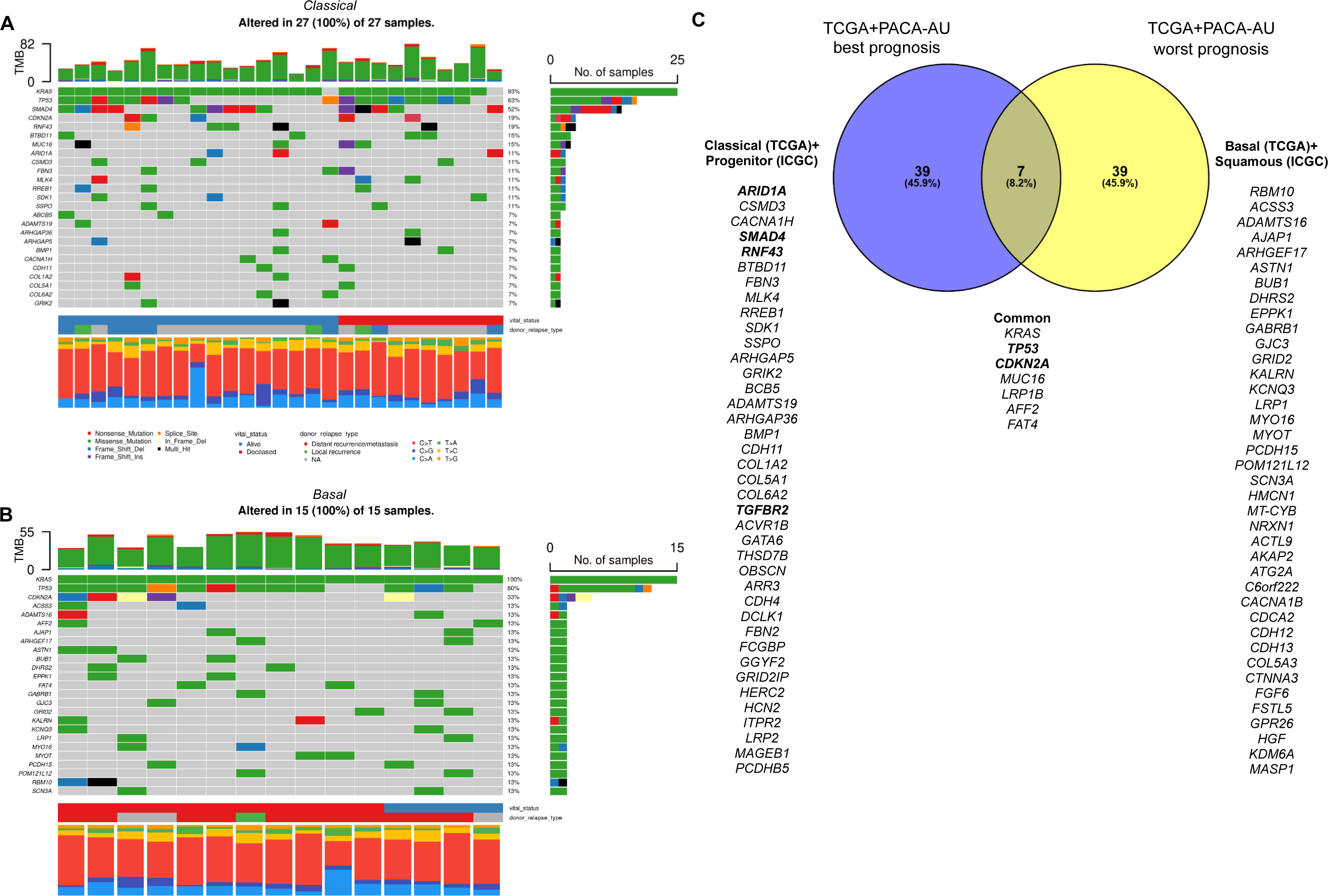
Transcriptomic stratification in PDAC reveals subtype-specific somatic variants. Oncoplots* of the top 25 most frequently mutated genes for consensus **(A)** n=27 *classical*-type and **(B)** n=16 *basal*-type PDAC cases (TCGA). **(C)** Overlap^ between the somatically mutated genes associated with best/worst prognosis subtypes across TCGA and ICGC PACA-AU cohorts combined. Genes highlighted in **bold** contain Tier 1 predicted oncogenic driver variants that have associated pharmacological inhibitors or chemotherapies (see Supplementary Table S1). *Mutated genes are ranked in order of the *total* number of *mutations* in each given gene (where genes may have >1 mutation present; black ‘multi-hit’), while the percentage to the right of each bar reflects the proportion of *samples* altered in the cohort. ^Created in [Venny 2.1](https://bioinfogp.cnb.csic.es/tools/venny/index.html).

### Identifying treatment biomarkers

*In silico* functional analysis of *all* patient-specific somatic variants identified, using the most recent, freely available embedded Cancer Genome Interpreter (CGI)(23) analytic tool, identified numerous biomarkers of response/resistance to existing clinical treatments and/or pharmacological inhibitors (Supplementary Table S2) in various cancer contexts, including multiple variants in *KRAS*, *TP53*, *CDKN2A* and *LRP1B*. Although *KRAS* is extensively mutated across both prognostic groups, *KRAS* p.G12C driver variants were unique to the best prognosis subtype tumours (Supplementary Table S2). This variant, rare in PDAC (<1% patients)(24), has been shown to preferentially drive the RAF/RAL pathway, while the more common *KRAS*^G12D^ mutation (∼30% PDAC patients) favours the PI3K/AKT pathway(24, 25). Targeted *KRAS*^G12C^ inhibitor sotorasib has recently received FDA approval for treatment of mNSCLC(26), and has been shown to be safe and effective in treatment of advanced mPDAC, in a Phase I/II trial in n=38 patients(27). Additionally, certain variants in *CDKN2A* (L104V, E120*, R58*, R80*) have been linked with treatment resistance to PD1 inhibitors, and treatment response to CDK4/6 inhibitors in cutaneous melanoma (CM; Supplementary Table S2), highlighting the usefulness of PGPA in identifying potentially clinically relevant patient- and cancer-specific therapeutic targets.

Of the somatic variants identified, CGI oncogenic classifications (bioactivity) (Supplementary Table S1), revealed several to be TIER 1 predicted driver variants, i.e. the gene activity is confirmed relevant to cancer, with mutations identified effecting oncogenic transformation. Only one was identified in the poorest outcome patient group – a splice donor variant in central cell-cycle regulator gene *ATM*, and linked with response to cisplatin chemotherapy, PARPi by olaparib, and PD1/PD-L1 inhibition in other solid tumours (Supplementary Table S2). However, this variant was uncommon in the sample set (1/16). Conversely, several TIER 1 oncogenic driver variants were identified in the best prognosis patient group, with some associated with response/resistance to specific drugs in other solid tumours, suggesting possible utility in pancreatic cancer: *ARID1A* (p.E1542*, p.Q1277*; responsive to EZH2, PD1, ATR & PARP inhibitors), *RNF43* (W159*, responsive to porcupine inhibitor) and *TP53* (S166*, Y205S, D259V, M246R, S241F, L194H, N131I; resistance to CDK4/6 inhibitor abemaciclib, cisplatin, MDM2 inhibitor; responsive to ATR inhibitor AZD6738, doxorubicin, decitabine, gemcitabine, mitomycin C).

Results of the above detailed output are also summarised in alluvial plots, showing clinically-actionable targets present in selectable proportions of the filtered dataset (5%-25%) and their responsiveness/resistance to available drugs (Supplementary Figure 2A, B), as well as a visual summary of the number of druggable gene categories represented in the selected dataset (Supplementary Figure 2C, D), that shows different clinically actionable genome targets between the two prognostic groups. While the most commonly mutated genes are common between prognostic subtypes (≥25% of patients from both groups contain similar *KRAS* and *TP53* variants), distinct biomarker/drug combinations are apparent when less common variations are considered: only one other gene in the poor prognosis group was identified as harbouring variants linked to (among others) responsiveness to small molecule AURKA-VEGF inhibitor ilorasertib (CDKN2A R58*), whereas the better prognosis subgroup is associated with 6 additional gene/biomarker candidates (*RNF43*, *PIK3CA*, *ERCC4*, *CTNNB1*, *CDKN2A*, *ARID1A*), that have shown promise in treating other solid tumours. Additionally, by exploring the available CCLE database of 60 pancreatic cell lines, *in vitro* models with/without *KRAS* and/or *TP53* variants may be identified to support downstream functional studies.

These results demonstrate the value of PGPA in contextualising individual patient genetic profiles in suggesting possible treatment options, or refining research areas to pursue for more effective, stratified approaches in pancreatic cancer.

### Characterising patterns of gene expression in best- and worst-outcome tumours

Inspecting the top 250 differentially expressed genes in both TCGA and ICGC filtered patient subgroups confirms scant overlap between genes differentially expressed in best and worst-outcome PDAC tumours. However, *classical/progenitor* tumours appear to have higher *TP53* expression compared to *QM/basal/squamous* tumours, consistent with its role as tumour suppressor (Figure 4A) and mirrored in ICGC *progenitor* and *squamous* patient subgroups(17) (Supplementary Figure 3). Subtype-specific differences in expression were also observed for *MUC16* (encoding CA125 membrane glycoprotein) which was over-expressed in *QM/basal/squamous* subtype tumours, compared to *classical/progenitor* cases (Figure 4B). Considering all n=402 confirmed PDAC in PACA-AU (the largest single transcriptomic dataset), PGPA shows that *MUC16* over-expression is associated with significantly reduced patient survival (logrank p=0.011; HR=2.23) (Figure 4C), where no association was found for neuroendocrine tumours (Figure 4D). Elevated CA125 has also recently been shown to be an independent prognostic marker of significantly shorter survival in n=207 resectable PDAC patients, both before and after treatment(28). Functionally, CA125 over-expression has been shown to promote tumourigenesis *in vitro* and *in vivo*(29, 30), and monoclonal antibody mAb AR9.6 has recently shown potential as a specific and effective inhibitor of CA125 and its oncogenic effects in pancreatic and ovarian cancers(30, 31).

**Figure 4.**
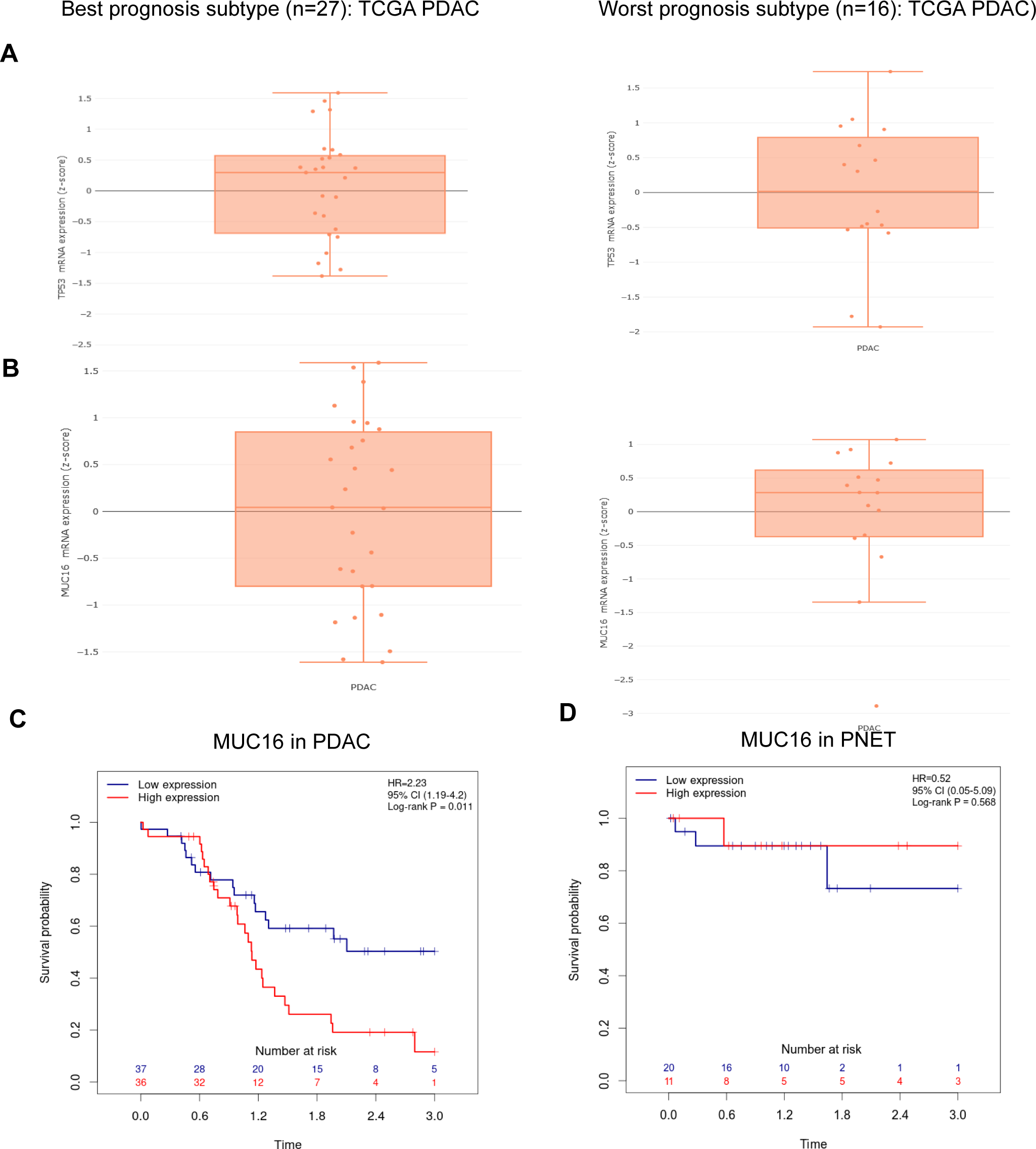
Differentially expressed genes between classical/progenitor and basal-like/QM/squamous TCGA PDAC tumours. Box plots showing the trends of **(A)** TP53 and **(B)** MUC16 mRNA expression levels across all patients in each filtered TCGA PDAC group; best prognosis (n=27; classical/progenitor; left) and worst prognosis (n=16; basal/squamous/QM; right). **C**. Kaplan-Meier curve showing elevated MUC16 expression significantly associated with lower patient survival over 3 years, from n=402 PACA-AU PDAC patients with expression and outcome data (logrank p=0.011; hazard ratio (HR)=2.23). **D.** No association between MUC16 mRNA expression levels and outcome were observed in n=65 neuroendocrine carcinomas (PAEN-AU).

### Identifying clinically actionable genomic alterations in KRAS wild-type PDAC tumours

Several studies have explored the genetic landscape of *KRAS* wild-type tumours, delineating several alterations that occur frequently in the absence of mutant *KRAS* (20, 32–34). In addition, a significant enrichment for somatic aberrations that target the RAS-MAPK pathway, either upstream or downstream of *KRAS*, has been observed in up to one-third of *KRAS* wild-type tumours(20, 34),where *BRAF* alterations were prevalent and mutually exclusive with *KRAS* mutations(34). However, alterations within genes that are not typically associated with *RAS* signalling have also been widely identified across *KRAS* wild-type PDAC tumours, and require further investigation to determine functional relevance(17, 20).

### Alternative oncogenic drivers amongst KRAS wild-type PDAC tumours

We used GENIE(10) as the largest available resource to identify n=756 *KRAS* wild-type PDAC samples. To identify other likely molecular drivers in these tumours, predictions from CGI were analysed to evaluate the distribution of altered genes and their associated pathways. Mutations were detected across several genes previously reported to be altered in *KRAS* wild-type PDAC cases, including *TP53* (mutated in >40% of the samples), *GNAS* and *BRAF*(35) (Figure 5A). *In silico* biomarker predictions also showed that *ARID1A*, *BRAF, CDKN2A, GNAS, PIK3CA and TP53* variants demonstrated therapeutic potential in response to PARP, tyrosine kinase and VEGF inhibitors, immunotherapies and several chemotherapies (Figure 5B).

**Figure 5.**
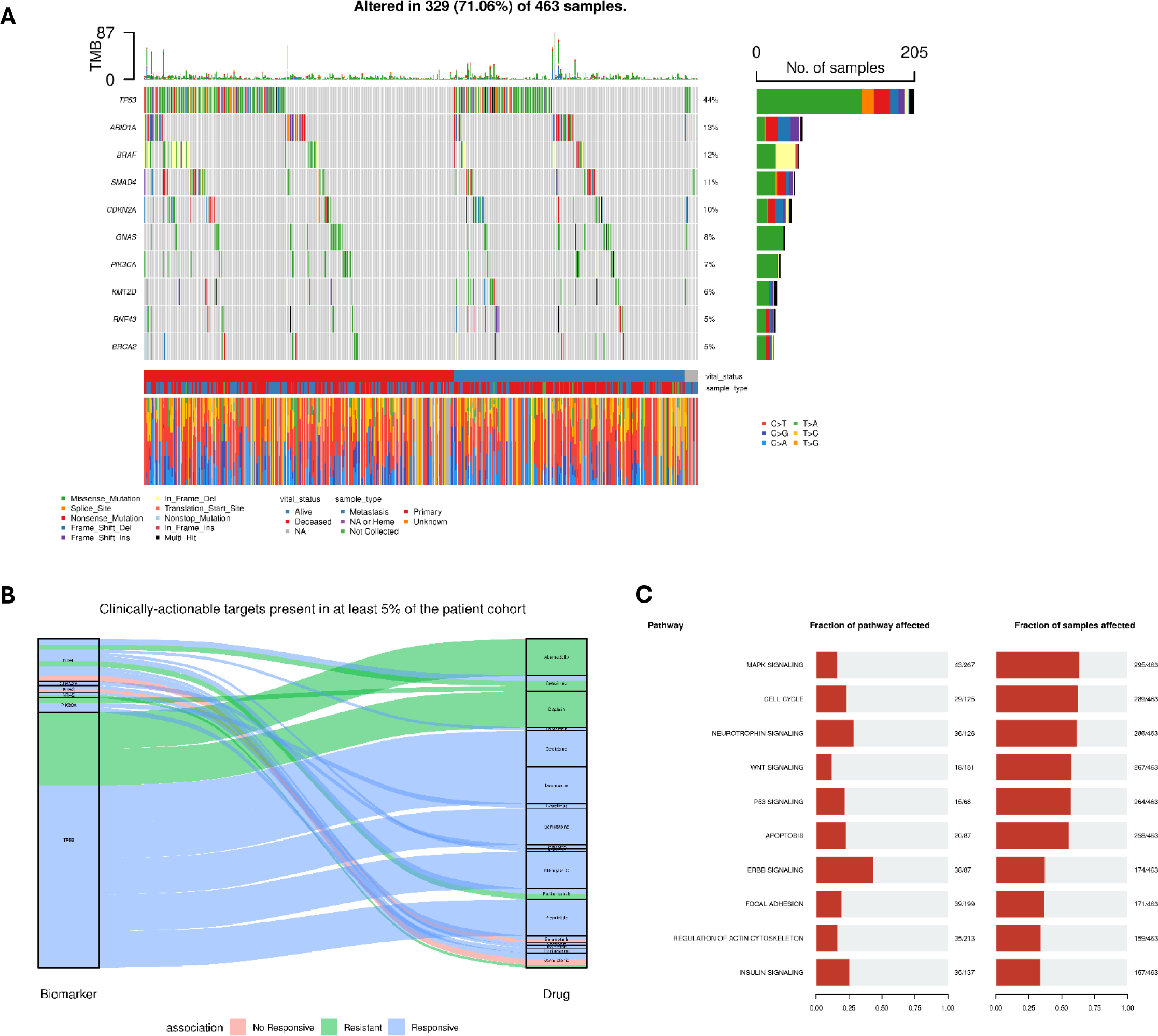
Frequently altered genes and biological pathways amongst n=756 *KRAS* wild-type PDAC tumours from GENIE. **(A)** Oncoplot showing the top 10 most frequently mutated genes in *KRAS* wild-type PDAC tumours (confirmed somatic missense mutations filtered out; insertions or duplications may still be present). **(B)** Alluvial plot showing gene targets harbouring any variants with therapeutic biomarker potential in ≥5% of patients, as identified by the Cancer Genome Interpreter and based on data from [OncoKB](https://www.oncokb.org/), [CIVic](https://civicdb.org/welcome) (Clinical Interpretation of Variants in Cancer) and the [Cancer Biomarkers database](https://www.cancergenomeinterpreter.org/biomarkers). **(C)** Altered biological pathways amongst *KRAS* wild-type PDAC tumours include MAPK and p53 signalling, as derived from the KEGG pathway database. The proportion of genes mutated in each pathway (left) and the proportion of all KRAS wild-type patients affected (x-axis) are given.

The analysis of altered signalling pathways amongst mutated genes across *KRAS* wild-type samples revealed frequent alterations in pathways associated with MAPK signalling, P53 signalling, neurotrophin, cell cycle, wnt and apoptosis signalling, consistent with previous characterisations of core biological pathways involved in PDAC development and progression(17, 18, 20, 29, 35). (Figure 5C). These findings highlight the utility of PGPA for the prioritisation of functionally and biologically relevant variants amongst subgroups of PDAC tumours, with important implications for the characterisation of distinct molecular pathologies and the identification of novel therapeutic opportunities.

Using the PGPA Analytics Hub, we demonstrate how our integrated high-performance visualisation and analysis tool can be used to investigate the link between genomic and transcriptomic features and phenotypes of pancreatic cancer, providing an important step in defining potential subtype-specific therapeutic vulnerabilities.

## The PCRFTB Data Return Module

The vision of precision medicine has driven unprecedented interest into biomarker-based studies (genomics, transcriptomics, proteomics) for pancreatic cancer, which are being adopted across research and development from early discovery through to clinical research and trials(36). Fundamental to biomarker research is access to quality biospecimens and samples that have been well annotated with clinical and molecular data(6). Whilst many biobanks have invested heavily in the IT infrastructure of sample management, most platforms are facing challenges in the effective sharing of returned data to drive investigative research across the pancreatic research community. A major challenge is the rise of so-called ‘big data’ from e.g. NGS and images that need to be integrated with large quantities of primary/secondary care information and other real-world healthcare data. Traditional biobanks are not usually set up to leverage these innovations, which have the potential to improve patient outcomes and accelerate the development and delivery of new therapies. PGPA is now the primary bioinformatics platform of the PCRFTB, providing a unique integrated resource of biological materials and associated clinical, molecular and radiological/imaging data.

In addition to providing a direct link to sample requests from the PCRFTB, the newly incorporated data return module in PGPA hosts clinical and molecular data returned to the biobank from studies undertaken using PCRFTB specimens, where published findings have been made available for researchers to review and analyse prior to submitting a tissue request. Studies are categorised according to the type of -omics data generated for each project (i.e., genomic, transcriptomic, proteomic), with alternative data types (e.g., summaries of staining or imaging results generated from experimental investigations) classified “other”, to simplify use. These classifications are presented in a summary table, which also provides a description of each project and details the different sample types (i.e., blood, tissues, cell lines) and cohort sizes used for each project. Users also have the option view this information as clinical summary plots for each individual cohort, prior to exploring available molecular data from each study.

Like other tissue repositories(37, 38), PCRFTB has implemented a data return policy, where anonymised data derived from banked samples is returned to the tissue bank on completion of the study and made freely available to the research community, regardless of whether the study is ultimately published(6). As associated sample datasets develop, more in-depth integrative analyses will be possible. Under *PCRFTB Data*, PGPA lists the details of any returned data and associated sample characteristics available for analysis, while *PCRFTB Research Projects* links directly to the relevant published report. So far, this has further enriched the data available for banked tissue sample and/or patients, and currently includes data on stromal(39), urinary miRNA(40), metallomic(41) and volatile organic compound(42) biomarkers, proteomics (ELISA)(43), circulating tumour cells (CTCs) and xenotransplantation models(44)(45), germline and somatic mutations(46), a phase I clinical trial(47), and risk(48) and recurrence(49) predictions incorporating electronic health record data. To date, PCRFTB has processed 38 Expressions of Interest, supported 33 research projects and 19 peer-reviewed publications. Ultimately, each study contributes to the development of a ‘digital fingerprint’ for each patient, linking multi-modal data with longitudinal clinical information.

PGPA Analytics Hub is the web-based portal through which this enriched dataset can be accessed and compared with large scale pancreatic -omics data, with the unique benefit of also providing access to additional patient samples (*via* PCRFTB) for subsequent validation of molecular alterations with clinical potential. Here, we demonstrate the added value of tissue banking to precision cancer medicine, to translate research findings into prognostic and therapeutic tools using well-annotated curated tissues and associated clinical data.

## Discussion

A rapid expansion in high-throughput genomic and transcriptomic profiling of pancreatic diseases necessitates the development of sophisticated yet user-friendly analytics hubs to host the growing compendium of molecular and clinical datasets and enable integrated mining and analysis of available results. PGPA supports a range of data modalities to enable users across the diverse international pancreatic research community to identify and investigate trends in molecular data across disparate cohorts of patients, samples and cell lines easily and effectively. PGPA also provides an unprecedented opportunity to characterise distinct variations in both tumour mutation landscapes and gene expression profiles that are associated with prognostic molecular subtypes. Candidate tumour drivers and biomarkers predictive of response to existing and novel clinical treatments can be identified and visualized, allowing suitable targets for downstream validation and pharmacological testing to be prioritised without the need for laborious data retrieval or processing tasks. Furthermore, PGPA is the gateway to a national tissue bank repository of >60,000 samples from >3,400 patients and a growing repository of digitised radiological and H&E images, that researchers can access independently and apply for donor samples that match with their research question. The selection of samples is significantly improved by the availability of high-quality clinical data, curated and maintained by PCRFTB.

Pancreas Genome Phenome Atlas is also the bioinformatics platform of the PCRFTB, pioneering a new generation in biobanking to support effective data sharing and promote collaborative studies, democratizing access to complex cancer genomics. By harmonising PCRFTB samples with clinical and molecular information from datasets returned to the biobank, PGPA provides an essential platform to support translational pancreatic research and fuel discoveries that can manifest clinically meaningful benefits for patients. PCRFTB recently launched internationally (https://www.pcrf.org.uk/news/tissue-bank-launches-internationally/), improving opportunities for high-quality research into earlier diagnosis and treatment of pancreatic cancer. As -omics driven research continues to drive efforts to advance the characterisation of pancreatic diseases, PGPA supports the ongoing data analysis, integration and visualisation needs of the growing research community.

## Availability of data and materials

PGPA relies on publicly available datasets from TCGA (doi: 10.1016/j.ccell.2017.07.007), GENIE doi: 10.1158/2159-8290.CD-17-0151), ICGC (doi: 10.1093/database/bar026, doi: 10.1038/s41587-019-0055-9 and CCLE (doi: 10.1038/nature11003). ).

## Supplementary Data

Supplementary Data are available at NAR Online

## Supporting information

Suppl. Table S1_bioactivity

Suppl. Table S2_biomarkers

## Data Availability

N/A - all data presented in the study are from publicly available resources (TCGA, ICGC, GENIE, CCLE).

https://portal.gdc.cancer.gov/

https://dcc.icgc.org/

https://sites.broadinstitute.org/ccle/datasets

https://www.aacr.org/professionals/research/aacr-project-genie/aacr-project-genie-data/

## Acknowledgements

PCRFTB group authors and recruitment site: Mo Abu Hilal (Southampton), Bilal Al Sarireh (Swansea), Somaiah Aroori (Plymouth), Ali Arshad (Southampton), Satyajit Bhattacharya (The London Clinic, London), Brian Davidson (Royal Free, London), John Isherwood (Leicester), Deep Malde (Leicester), Stuart Robinson (Newcastle), Zahir Soonawalla (Oxford). We are especially grateful to the patients and their families who have donated samples and time to support this research.

## Ethics approval

All donors provide written, informed consent, and all samples are collected, processed and stored at each of the participating centres (Barts, Leicester, Swansea, Oxford, Royal Free (London), Southampton, Newcastle, Plymouth, The London Clinic) under one Research Ethics Committee reference (13/SC/0593, renewed 18/SC/0629, renewed 23/SC/0282).

## Consent for publication

Not applicable

## Author contributions

Study conception & design: CC, JO, AZMDU; Data acquisition: AZMDU, TSK, EG, GJT, MA, AI, AS, RR, CH, PCRFTB; Data analysis & interpretation: LS, HRA; software creation: JO, CC; writing (drafting): LS (substantial revision): HRA; Funding: HMK, CC. All authors read and approved the manuscript.

